# Neighbourhood characteristics associated with the geographic variation in laboratory confirmed COVID-19 in Ontario, Canada: a multilevel analysis

**DOI:** 10.1101/2021.04.06.21254988

**Authors:** Tristan Watson, Jeffrey C. Kwong, Kathy Kornas, Sharmistha Mishra, Laura C. Rosella

**Author notes:** **CORRESPONDING AUTHOR:** Dr. Laura C. Rosella, Dalla Lana School of Public Health, Health Sciences Building, 6^th^ floor, 155 College Street, Toronto, Ontario, M5T 3M7.

## Abstract

**Purpose:** There is limited information on the role of individual- and neighbourhood-level characteristics in explaining the geographic variation in the novel coronavirus 2019 (COVID-19) between regions. This study quantified the magnitude of the variation in COVID-19 rates between neighbourhoods in Ontario, Canada, and examined the extent to which neighbourhood-level differences are explained by census-based neighbourhood measures, after adjusting for individual-level covariates (i.e., age, sex, and chronic conditions).

**Methods:** We conducted a multilevel population-based study of individuals nested within neighbourhoods. COVID-19 laboratory testing data were obtained from a centralized laboratory database and linked to health-administrative data. The median rate ratio and the variance partition coefficient were used to quantify the magnitude of the neighbourhood-level characteristics on the variation of COVID-19 rates.

**Results:** The unadjusted median rate ratio for the between-neighbourhood variation in COVID-19 was 2.22. In the fully adjusted regression models, the individual- and neighbourhood-level covariates accounted for about 44% of the variation in COVID-19 between neighbourhoods, with 43% attributable to neighbourhood-level census-based characteristics.

**Conclusion:** Neighbourhood-level characteristics could explain almost half of the observed geographic variation in COVID-19. Understanding how neighbourhood-level characteristics influence COVID-19 rates can support jurisdictions in creating effective and equitable intervention strategies.

## INTRODUCTION

The spread of the coronavirus disease 2019 (COVID-19) remains a significant public health problem in many jurisdictions, including North America. In March 2020, the province of Ontario, Canada, enacted the “Emergency Management and Civil Protection Act” in order to help contain the spread of the COVID-19 [1]. This included interventions such as closing non-essential services, regional restrictions or requirements and developing case finding and management protocols. Despite these public health interventions, there have been growing concerns and reports that the spread of COVID-19 disproportionately affects marginalized and vulnerable groups [2–10].

The role of socio-demographic factors and geography of residence on the risk of acquiring COVID-19 has been examined in an increasing number of studies. In the USA, a study focused on county-level comparisons of COVID-19 cases found that counties with a higher proportion of black people had more COVID-19 diagnoses [11]. A recent Canadian report found that people living in marginalized neighbourhoods were more likely to test positive for COVID-19 [12]. Another Canadian study found that COVID-19 diagnoses were more common in black and immigrant communities in Canada [6]. A common feature in these studies is the lack of available individual-level data on important socio-demographic variables (e.g., occupation, income, housing, transportation, and physical environment). Due to the lack of individual-level data on social determinants of health, researchers have been using geographically aggregated data from sources, such as census data, or geocoded to a postal code [6,9,11,13].

There has been a lack of studies that have quantified the magnitude of the general contextual effect (i.e., effect of cluster level on outcome) of neighbourhood-level characteristics on the variation of COVID-19 rates while including individual-level variables. Most of the COVID-19 studies have been at only the individual- or ecological-level. There is a paucity of multilevel analyses that were conducted to understand the characteristics of infection risk across various communities. Multilevel analyses are important for many diseases because there is consensus that neighbourhood-level interventions (e.g., local access to of testing and mask-wearing policies) and characteristics can contribute to individual-level health outcomes [13,14]. By improving our understanding of how neighbourhood-level characteristics influence COVID-19 rates, jurisdictions can be better prepared to create effective and equitable intervention strategies [15].

The objective of this study was to quantify the magnitude of the variation in COVID-19 rates across neighbourhoods in Ontario, Canada, and to examine the extent to which neighbourhood-level differences are explained by the census-based neighbourhood measures after adjusting for individual-level covariates (i.e., age, sex, and select chronic conditions).

## METHODS

### Context and setting

This study was conducted using population-based data from the province of Ontario, Canada. Ontario is ethnically diverse, with an estimated 29.1% of the population comprised of immigrants as of 2016 [16]. All residents of Ontario are eligible for universal health coverage. COVID-19 interventions that were in place in Ontario during the study period included case management, temporary closure of schools, temporary closure of non-essential services, recommendations to work from home, mandatory use of face coverings, implementation of virtual care, and mandatory 14-day quarantine for international travellers [1].

### Analytical Cohort / Study Cohort Participants

The analytic cohort was created through the linkage of multiple population and health administrative databases held within ICES. These datasets were linked using unique encoded identifiers and analyzed at ICES. ICES is an independent, non-profit research institute whose legal status under Ontario’s health information privacy law allows it to collect and analyze health care and demographic data, without consent, for health system evaluation and improvement. Further details about the data sources are included in **Appendix A**.**1**. The COVID-19 laboratory results were obtained using polymerase chain reaction (PCR) test during the observation period from the Ontario Laboratories Information System (OLIS) database. The analytic cohort includes all publicly insured Ontarians, with a positive COVID-19 test, aged 20 years or older, with a valid postal code, not living in a long-term care home, alive on March 1, 2020, and included events up to July 31, 2020 (N = 26,397) (**Figure 1**). If a person tested positive for COVID-19 multiple times in the observation window, the first positive test data was used. The focus of this study is on COVID-19 cases in the community-dwelling population. Therefore, people with a recent stay in a long-term care (LTC) facility were excluded because the community-based spread of infection is different from the spread of infection in LTC facilities [17,18].

**Figure 1.**
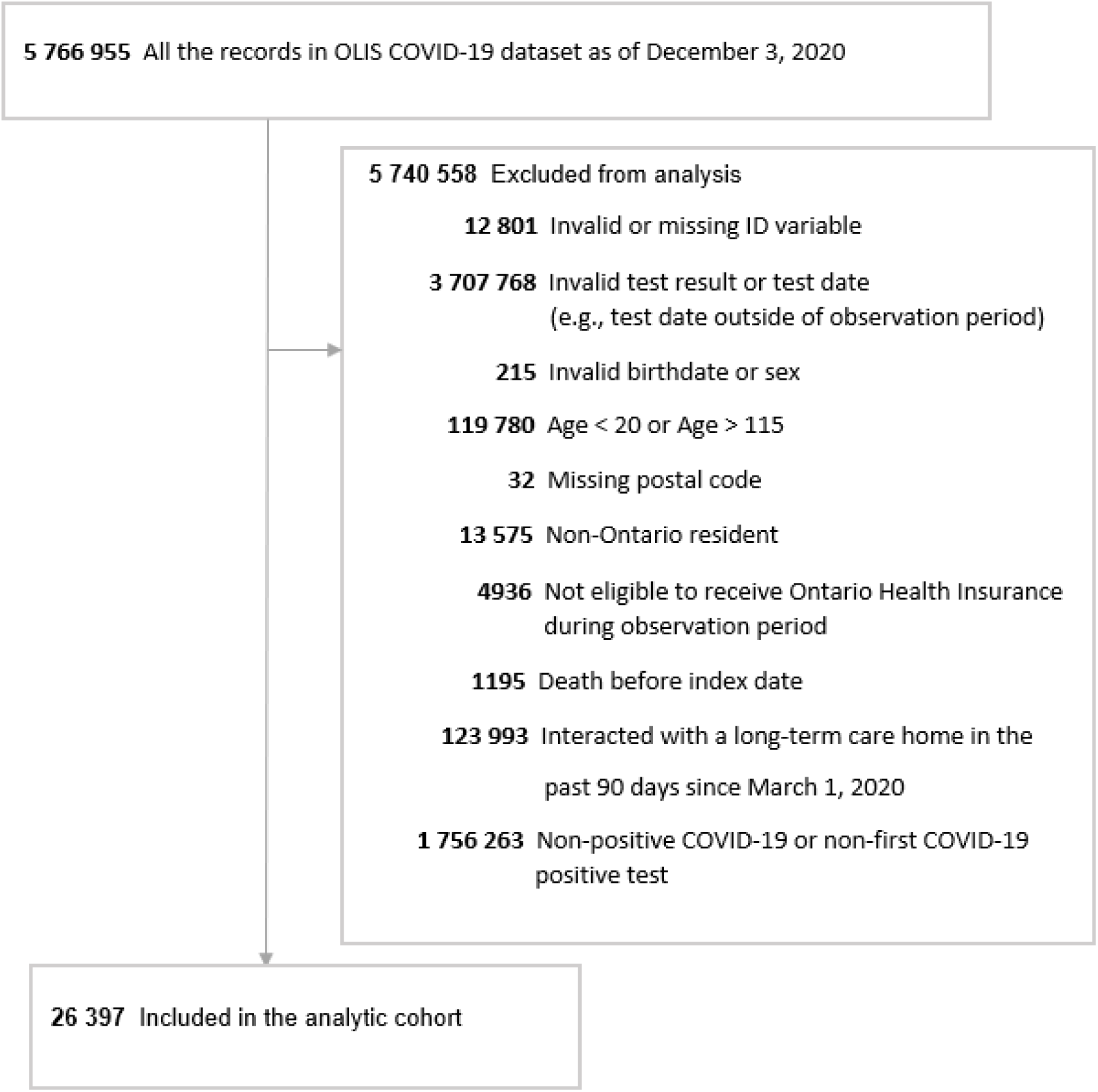
Study Cohort Flow Diagram. Abbreviation: OLIS, Ontario Laboratories Information System.

### Measures

#### Outcome

The primary outcome, defined a priori, was the neighbourhood rate of laboratory-confirmed COVID-19 per 1000 people in each neighbourhood in Ontario, Canada. The various neighbourhood rates of COVID-19 were calculated using the number of positive laboratory-confirmed COVID-19 tests as the numerator and total number of people at risk in each neighbourhood at start of the study period divided by 1000 as the denominator.

#### Exposures

Data were structured in a two-level nested hierarchical structure: individuals who tested positive for COVID-19, and neighbourhoods. Exposure variables were examined at the individual-level and the neighbourhood-level.

Individual-level variables included age (age 20-34, 35-49, 50-64, 65-114), sex (male or female), history of asthma (yes or no), history of diabetes (yes or no), history of hypertension (yes or no), history of congestive heart failure (CHF) (yes or no), and history of chronic obstructive pulmonary disease (COPD) (yes or no). Age categorizations were based on the quartiles of the age distribution for our study population. These variables have been included in this study because they are associated with different rates of being tested and testing positive for COVID-19 [12]. For example, people with chronic conditions may be tested more frequently for COVID-19 due to more interactions with the healthcare system. For all chronic conditions, the ascertainment of each chronic condition was based on physician diagnosis codes and the look-back period included all available data before March 1, 2019. Further details about the diagnostic codes used for the ascertainment of chronic conditions are in **Appendix Table A**.**1**.

The census-based neighbourhood measures were derived from the 2016 Ontario Census Area profiles summarized at the forward sortation area (FSA). In Canada, the FSA represents a geographic region based on the first three characters in a Canadian postal code. In 2016, Ontario had 523 FSAs represented in the 2016 Census. In this present study, neighbourhoods are operationalized as FSAs. The FSA is a stable standard geographic area used for linkage with the Census data. The following four considerations guided the choice and development of census-based neighbourhood-level variables in this study: (1) a priori knowledge of potential conditions that increase potential virus exposure, (2) influential covariates common in past infectious disease epidemiology studies, (3) area-based measures that might be amenable to public health or policy intervention, (4) and area-based measures that are representative and not collinear based on a cluster analysis [6,13]. Five census-based neighbourhood-level variables were generated that met these criteria: the average number of people living in a home, proportion of people working a sales job, the proportion of people working a managerial job, the proportion of people that drive to work, and the median after-tax income. Further details about the definitions of each Census variable used are in **Appendix Table A**.**2**.

### Statistical Analysis

Three multilevel Poisson regression models were used to evaluate the contribution of the census-based neighbourhood measures to the neighbourhood rates of COVID-19. Variables were added sequentially to the three different multilevel Poisson regression models. First, a null model was fit (model 1), which only included neighbourhood-specific random intercepts. Second, a multilevel Poisson regression model was fit (model 2) with neighbourhood-specific random intercepts that added seven individual-level variables: age, sex, asthma, diabetes, hypertension, CHF, and COPD. Third, a multilevel Poisson regression model was fit (model 3) with neighbourhood-specific random intercepts that added five census-based neighbourhood-level variables.

For each of the three models, the Ontario population at risk of disease was used as the offset variable. The census-based neighbourhood-level measures were standardized to have a mean of zero and a standard deviation of 1. Before fitting these regression models, observations from the cohort were removed if their FSA contained fewer than 20 people or had a missing covariate (n = 57). The final analytical cohort had 26,340 COVID-19 cases. The cohort was aggregated across the different covariates (i.e., for each FSA, by age group, sex, and chronic disease flags) in the model, with each stratum in the dataset representing a unique combination of the covariates. People residing in the same FSA had the same census-based variables. For each of the five census-based variables, the assumption of linearity was assessed using the fully adjusted model (i.e., model 3), by using a Wald chi-squared test based on the use of restricted cubic splines [19]. The Wald-chi square test (p > 0.05) showed that that there was a linear relationship between each census-based continuous variable and the COVID-19 rates per 1000 in the fully adjusted model.

The neighbourhood-level variance and median rate ratio (MRR) for neighbourhood-level effects were calculated for each model. The MRR is the median value of the rate ratio that would have been obtained in the outcome (i.e., rate of COVID-19) if we took repeated random samples of two individuals with identical covariates from two different neighbourhoods (high-risk versus low-risk neighbourhood) [20]. The magnitude of the MRR reflects the between-neighbourhood variation in the neighbourhood rate of COVID-19. By using a summary measure of the between-neighbourhood variation, it provides an overall summary of neighbourhood effects without the need for comparing individual neighbourhoods against a single reference neighbourhood.

The proportional change of the variance (PCV) was calculated for models 2 and 3 in reference to model 1 [21,22]. The PCV quantifies how much of the neighbourhood differences in COVID-19 rates may be attributable to individual-level or neighbourhood-level differences based on covariates included and not included in our study.

The variance partition coefficient (VPC) was calculated for model 3. The VPC is a measure of the proportion of the total observed individual variation in the outcome due to between-cluster variation [20]. The VPC ranges from 0 to 1, where one indicates that all the variation in the outcome is due to between-cluster variation. A larger VPC indicates a larger cluster effect (e.g., neighbourhood effect). In other words, individuals from the same neighbourhood are likely to have the same outcome. The VPC is equivalent to the intraclass correlation coefficient (because the data used in this study has a basic two-level hierarchical nested structure [20,21]. The VPC was only calculated for the different patient profiles based on typical covariate patterns and values of the offset variable. The VPC was evaluated at values ranging from 0.001 per 1000 to 0.277 per 1000 people, in increments of one. This represented the tenth percentile and 90^th^ percentile in the offset term, respectively, for the total population for each FSA, by age group, sex, and chronic disease flags. The examination of a range of offset variables shows the variation of the VPC across different population sizes, rather than relying only on the mean population size.

The structure of the random effects was assumed to follow a variance components structure. The multilevel regression models were run using the Gauss-Hermite quadrature technique with empirical sandwich estimates of the standard error that are robust to misspecification of the covariance structure [23–25]. Model fit statistics were produced for each model. SAS Enterprise Guide v.8.15 (SAS Institute Inc, Cary, NC) was used to create the data set and conduct the multilevel regression analyses. Stata/MP v15 (StataCorp, College Station, TX) was used to conduct the ICC-VPC analyses.

## RESULTS

### Descriptive statistics

A total of 26,397 patients with laboratory-confirmed COVID-19 between March 1 and July 31, 2020 were included (median age, 48 years, [interquartile range {IQR}, 33-60; range, 20-104 years]; 53% female) in the analytical cohort (**Table 1**). **Table 1** shows the demographic, chronic disease, and census-based neighbourhood-level characteristics of the analytical cohort. Of the five chronic diseases identified, hypertension (29% [95% CI, 28%-29%]) and diabetes (16%, [95% CI, 16%-17%) were the most prevalent. Among the census-based neighbourhood-level measures, the median (IQR) of the average household occupants in each neighbourhood was 3 (2-3). The median (IQR) percentage of households that drove as their main mode of commuting in each neighbourhood was 73% (57% - 80%). The median (IQR) of the median after tax household income in 2015 in each neighbourhood was $29,245 ($24,908 - $32,865) Canadian dollars. The median (IQR) for the proportion of workers in sales and managerial jobs in each neighbourhood were 24% (22%-27%) and 10% (8%-13%), respectively.

**Table 1.** Baseline demographic, chronic conditions, and census-based area characteristics of patients that tested positive for COVID-19 in Ontario, Canada between March 1, 2020 to July 31, 2020

|  | No. (%) |
| --- | --- |
| **Demographic** |  |
| Total No. | 26 397 |
| Age, median (IQR) [range], y | 48 (33-60) [20-104] |
| Age, y |  |
| 20-34 | 7225 (27) |
| 35-49 | 6839 (26) |
| 50-64 | 7669 (29) |
| 65-114 | 4664 (18) |
| Sex |  |
| Female | 13 954 (53) |
| Male | 12 443 (47) |
| **Chronic Conditions** |  |
| Diabetes | 4322 (16) |
| Hypertension | 7563 (29) |
| COPD | 1732 (7) |
| CHF | 799 (3) |
| Asthma | 3804 (14) |
| **Census-based Area** |  |
| Average Household Occupants, median (IQR) [range], No. | 3 (2-3) [1-4] |
| Percentage of Households driving, median (IQR) [range], % | 73 (57-80) [11-95] |
| Percentage of Sales / Service Jobs, median (IQR) [range], % | 24 (22-27) [12-43] |
| Percentage of Managerial / Professional Jobs, median (IQR) [range], % | 10 (8-13) [4-26] |
| Median After-tax Household Income (after-tax), median (IQR) [range], CAD \$ | 29 245 (24 908 – 32 865) [18 947-53 376] |

Abbreviations: COPD, chronic obstructive pulmonary disease; CHF, congestive heart failure, CAD, Canadian
Note: Definitions for the chronic conditions and census-based area characteristics are provided in Appendix Table A.1 and Appendix Table A.2.

### Multilevel regression analyses

Table 2 shows the results from three multilevel Poisson regression models sequentially adjusted for demographic characteristics, chronic conditions, and census-based neighbourhood-level characteristics. In the null multilevel Poisson regression model (model 1), the estimated mean intercept was 0.3503, while the variance of the neighbourhood-specific random effects was estimated to be 0.70. The MRR was 2.22. After adjusting for the individual-level (i.e., age, sex, and chronic conditions) characteristics (model 2), the estimated mean intercept was −0.1865., while the variance of the neighbourhood-specific random effects was estimated to be 0.69. In model 2, the PCV was 1%. Therefore, 1% of the neighbourhood variation in COVID-19 rates in the null model was attributable to age, sex, and chronic condition characteristics. After adjusting for both individual- and neighbourhood-level characteristics (model 3), the estimated mean intercept was −0.1117, while the variance of the neighbourhood-specific random effects was estimated to be 0.39. In model 3, the PCV was 44%, which means an additional 43% of neighbourhood variation in COVID-19 rates in the null model was attributable to the five census-based neighbourhood characteristics.

**Table 2.** Sequential multilevel Poisson count regression models for COVID-19 for patients with a laboratory-confirmed positive test for COVID-19 in Ontario, Canada between March 1 and July 31, 2020

|  |  |  |  |
| --- | --- | --- | --- |
| Neighbourhoods (i.e., FSA) = 513<br>Individuals = 26 340 | **Model 1 (Null model)** | **Model 2 (model 1 + age, sex, and chronic conditions)** | **Model 3 (model 2 + census-based neighbourhood measures)** |
| Mean rate of COVID-19 per 1000 people (intercept) | 0.35 | -0.19 | -0.11 |
| **Individual Level** | **RR (95% CI)** | **RR (95% CI)** | **RR (95% CI)** |
| Age, yr (ref. 65-114) |  |  |  |
| 20-34 |  | 1.68 (1.53-1.85) | 1.68 (1.53-1.85) |
| 35-49 |  | 1.54 (1.41-1.68) | 1.54 (1.41-1.68) |
| 50-64 |  | 1.57 (1.46-1.69) | 1.57 (1.46-1.68) |
| Sex (ref. men) |  | 1.10 (1.04-1.16) | 1.10 (1.03-1.16) |
| Diabetes |  | 1.36 (1.30-1.42) | 1.35 (1.29-1.42) |
| Hypertension |  | 1.30 (1.24-1.36) | 1.30 (1.24-1.36) |
| COPD |  | 0.99 (0.93-1.06) | 1.00 (0.93-1.06) |
| CHF |  | 1.62 (1.49-1.77) | 1.62 (1.49-1.77) |
| Asthma |  | 0.98 (0.92-1.03) | 0.98 (0.92-1.03) |
| **Census-based Neighbourhood Level** | **RR (95% CI)** | **RR (95% CI)** | **RR (95% CI)** |
| Average number of household occupants |  |  | 1.61 (1.50-1.72) |
| Proportion of households driving |  |  | 0.63 (0.59-0.67) |
| Proportion of people in sales / service Jobs |  |  | 1.07 (0.99-1.17) |
| Proportion of people in managerial / professional Jobs |  |  | 0.93 (0.86-1.01) |
| Median after-tax income in 2015 |  |  | 0.99 (0.91-1.09) |
| **Level 2 (Neighbourhood) Variance** | 0.7 | 0.69 | 0.39 |
| Proportional change in variance (PCV) by the new model | Reference | 1% | 44% |
| Median Rate Ratio for neighbourhood effects | 2.22 | 2.21 | 1.82 |
| **Model Fit Statistics** |  |  |  |
| Fit Statistics (-2LL) | 56475.40 | 55365.35 | 55106.16 |
| Pearson Chi-Square / DF | 1.28 | 1.01 | 1.02 |

Abbreviations: FSA, forward sortation areas; RR, rate ratio; CI, confidence interval; yr, year; ref., reference; COPD, chronic obstructive pulmonary disease; CHF, congestive heart failure.
Note: The neighbourhood was defined according to forward sortation areas (FSA). In Canada, the FSA represents a geographic region based on the first three characters in a Canadian postal code. Before fitting these regression models, observations from initial the cohort were removed if their FSA contained less than 20 people or had a missing covariate (n = 57). The final analytical cohort had 26,340 COVID-19 cases. The census-based neighbourhood level measures were standardized to a mean of 0 and standard deviation of 1. The patients in this
study had a laboratory confirmed positive test for COVID-19 in Ontario, Canada between March 1, 2020, and July 31, 2020.
Definitions for the chronic conditions and census-based area characteristics are provided in Appendix Table A.1 and Table A.2.

The MRR from a high-risk versus low-risk neighbourhood decreased proportionately, from an unadjusted MRR of 2.22 to 1.82 in the fully adjusted model. In the fully adjusted model, the median effect of clustering on the COVID-19 rate between neighbourhoods was greater in magnitude than effects for all the other examined individual-level and neighbourhood-level covariates because there was not a rate ratio outside of the final MRR (1.82) or the reciprocal of the MRR (1/1.82 = 0.549). This suggests that the neighbourhood clustering effect has a strong effect on the COVID-19 rate.

The VPC ranged from 0.001 when the size of the population at risk was 1 to 0.13 when the population at risk was 277 (or 0.277 per 1000) in the reference patient (**Figure 2**). Among individuals (male and 65-114 years old) who belong to a stratum with at least 277 people, 13% of the variation in COVID-19 rates was due to systematic differences between neighbourhoods, while the remaining 87% was due to within-neighbourhood between-subject differences. In the younger age groups, compared to the 65-114 age category, a greater proportion (∼ 20%) of the variation in COVID-19 rates was due to systematic differences between neighbourhoods.

**Figure 2.**
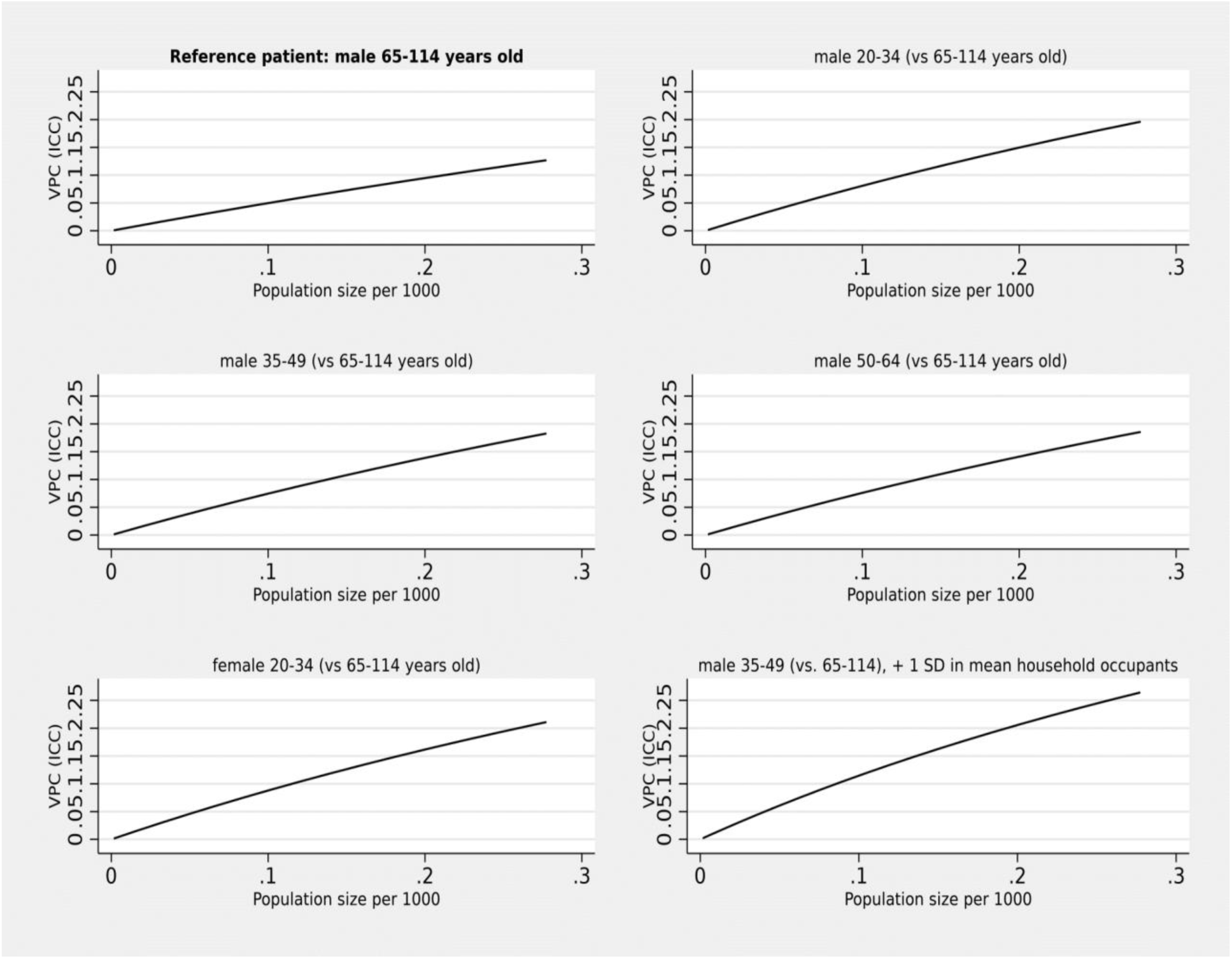
Variance partition coefficient (VPC) (intraclass correlation coefficient [ICC]) for six different patient profiles. Abbreviations: vs, versus; SD, standard deviation.

## DISCUSSION

Using a multilevel study, we found that individual- and neighbourhood-level factors combined accounted for 44% of the variation in COVID-19 rates between neighbourhoods. Nearly all this variation (43% overall) was attributable to neighbourhood-level characteristics examined in our study and only 1% was attributable to individual-level characteristics (i.e., age, sex, and selected chronic conditions).

In our study, neighbourhoods with a higher number of people living per dwelling were associated with higher COVID-19 diagnoses. Previous research has also shown that household crowding increases vulnerability to COVID-19 infection [26–28]. Public health interventions with the potential to support individuals who live in crowded housing and test positive for COVID-19 could include, in the short-term, the establishment of and community-mobilization for volunteer isolation centres and wrap-around quarantine support in the context of challenges to effective and safe isolation/quarantine at home. These measures should be considered within broader policy measures that act on structural determinants associated with overcrowded households, such as improving housing affordability [4]. Our study also showed associations with neighbourhood employment factors on neighbourhood variation in COVID-19 rates. Occupational factors can impact exposure to COVID-19 infection. For example, people working in essential industries may work in close proximity to others in the workplace and have more frequent exposure to infection [29]. Our findings underscore the importance of occupational factors on COVID-19 neighbourhood disparities and a need for interventions that support a healthy workforce, including personal protective equipment, detailed case management tracking, and paid sick leave.

This study has several strengths. We accounted for correlations within the data due to clustering with a multilevel model. By using a count-based multilevel model, the VPC can reveal how differences in population size alter the amount of variance explained in the model. By quantifying the contextual effects of neighbourhood on disparities in COVID-19 rates, this provide evidence of structural and neighbourhood-level factors that could be targets for policies and public health intervention.

Limitations include the potential for misclassification of neighbourhood measures as some neighbourhood features might have changed since 2016. The people with positive tests in our cohort may not be representative of Ontario because the population was not tested at random. Early in the pandemic, there was a bias towards testing symptomatic patients and people working in certain occupations (e.g., health care workers). This could lead to a type of selection bias known as collider bias (or detection bias), where some segments of the population are over or underrepresented in the analytic cohort [30–32]. By conditioning the results on those who were tested and symptomatic, this could lead to an over- or under-estimation of the MRR. In addition, about 15% cases of COVID-19 captured were not captured in the OLIS; therefore, our study under reports the number COVID-19 cases in the observation window compared to the numbers officially reported. However, given that most cases were captured in our analysis and missing cases don’t appear to cluster in a few select neighbourhoods, we do not expect that the missing cases would affect the MRR results in a meaningful way.

This study shows that socio-demographic neighbourhood-level factors explain almost half of the observed geographic variation in COVID-19 rates in Ontario, Canada. The research highlights the importance of neighbourhood-level characteristics in explaining the geographic variation in COVID-19 rates and suggests a need to address structural and social determinants within neighbourhoods to reduce COVID-19 disparities.

## Data Availability

The dataset from this study is held securely in coded form at ICES. While legal data sharing agreements between ICES and data providers (e.g., healthcare organizations and government) prohibit ICES from making the dataset publicly available, access may be granted to those who meet pre-specified criteria for confidential access, available at www.ices.on.ca/DAS

## List of Abbreviations

CHF: congestive heart failure
COPD: chronic obstructive pulmonary disease
FSA: forward sortation area
ICC: intraclass correlation coefficient
IQR: interquartile range
LTC: long-term care
MRR: median rate ratio
OLIS: Ontario Laboratories Information System database
PCR: polymerase chain reaction
PCV: proportional change of the variance
VPC: variance partition coefficient

## ACKNOWLEDGMENTS

This study was supported by ICES, which is funded by an annual grant from the Ontario Ministry of Health (MOH) and the Ministry of Long-Term Care (MLTC). The opinions, results and conclusions reported in this paper are those of the authors and are independent from the funding sources. No endorsement by ICES or the MOH is intended or should be inferred. Parts of this material are based on data and/or information compiled and provided by MOH and the Canadian Institute for Health Information (CIHI). However, the analyses, conclusions, opinions, and statements expressed herein are those of the authors, and not necessarily those of MOH and CIHI.

## ETHICS APPROVAL

This study received ethics approval from the University of Toronto’s Health Sciences Research Ethics Board. (Protocol #39356)

## AUTHOR STATEMENT

**Tristan Watson:** Conceptualization, Methodology, Formal analysis, Writing and Reviewing – Original Draft

**Jeffrey C. Kwong:** Methodology, Writing – review & editing

**Kathy Kornas:** Writing and Reviewing – original draft preparation

**Sharmistha Mishra:** Methodology, Writing – review & editing

**Laura C. Rosella:** Conceptualization, Methodology, Writing – review & editing, Supervision

