## Appendix A for "Neighbourhood characteristics associated with the geographic variation in laboratory confirmed COVID-19 in Ontario, Canada: a multilevel analysis"

**A.1** Data Sources

**Table A.1** Definitions of Physician-diagnosed Chronic Conditions

**Table A.2.** Definitions of Census-Based Neighbourhood Characteristics**:** constructs**,** statistical units, and operational definitions, using the 2016 (Ontario-specific) Canadian census data

### A.1. Data Sources

This study used population-based demographic and health administrative databases linked and analyzed at ICES in Toronto, Ontario. The COVID-19 laboratory results are from the Ontario Laboratories Information System (OLIS) database, which identifies individuals that were tested for COVID-19 in our study. The OLIS database contains information on the sex, birth date, specimen collection date, COVID-19 test result, and other variables. [12] OLIS does not capture all cases of COVID-19 in Ontario because some laboratories did not contribute results during the study period.

The OLIS data was linked to the Ontario Registered Persons databases (RPDB) using unique, encrypted Ontario Health Insurance Plan (OHIP) health card numbers. The RPDB is a central population registry file that permits linkage with data holdings held at ICES, and contains demographic information on birth date, sex, residential postal code, and other variables.

The health administrative databases include the OHIP claims database, Canadian Institute for Health Information Discharge Abstract Database (DAD), Continuing Care Reporting System (CCRS), Ontario Drug Benefit (ODB) claims database. Several disease-specific registries were utilized that contain regularly updated population cohorts and were created by applying validated data algorithms to the DAD and OHIP, specifically: Ontario Asthma Database, Ontario Congestive Heart Failure Database, Ontario Chronic Obstructive Pulmonary Disease Database, Ontario Hypertension Database, and the Ontario Diabetes Database (for more information, see eTable 1 in the Online Supplement). The OHIP, CCRS, and ODB databases were used to identify patients who interacted with the long-term care home 90 days prior to March 1, 2020.

### Table A.1: Definitions of Physician-diagnosed Chronic Conditions

| Chronic Condition | Case Definition/Reference | Database (latest data available) / *Notes |
|---|---|---|
| Asthma | **Case Definition**<br>≥1 hospitalization (from DAD) or ≥2 physician billings (from OHIP) within a two-year period with an asthma diagnosis code<br><br>OHIP<br>OHIP physician diagnostic code: 493<br><br>DAD<br>ICD-9 diagnostic code: 493<br>ICD-10 diagnostic codes: J45, J46<br><br>**Reference**<br>Gershon AS, Wang C, Guan J, Vasilevska-Ristovska J, Cicutto L, To T. Identifying patients with physician-diagnosed asthma in health administrative databases. Can Respir J 2009; 16: 183-8. | **Database**<br>Ontario Asthma Database (ASTHMA)<br><br>(March 31, 2019) |
| Chronic obstructive pulmonary disorder | **Case Definition**<br>≥1 hospitalization or ≥1 physician billing within a two-year period with a COPD diagnosis code<br><br>OHIP<br>OHIP physician diagnostic codes: 491, 492, 496<br><br>DAD<br>ICD-9 diagnostic codes: 491, 492, 496<br>ICD-10 diagnostic codes: J41, J42, J43, J44<br><br>**Reference**<br>Gershon AS, Wang C, Guan J, Vasilevska-Ristovska J, Cicutto L, To T. Identifying individuals with physician diagnosed COPD in health administrative databases. COPD. 2009; 6(5):388–94. | **Database**<br>Ontario Chronic Obstructive Pulmonary Disease (COPD)<br><br>(March 31, 2019)<br><br>* The COPD database is limited to adults age 35 and older. |
| Congestive heart failure | **Case Definition**<br>≥1 hospitalization (from DAD, SDS, or OMHRS) or ≥1 physician billing (from OHIP/ NACRS) followed by ≥1 hospitalization (from DAD)/emergency department visit (from NACRS) /physician billing (from OHIP) within a one-year period with a congestive heart failure diagnosis code<br><br>OHIP:<br>OHIP physician diagnostic code: 428<br>OHIP physician fee code: Q050<br><br>DAD, SDS, OMHRS, NACRS:<br>ICD-9 diagnostic code: 428<br>ICD-10 diagnostic codes: I500, I501, I509<br><br>**Reference**<br>Schultz SE, Rothwell DM, Chen Z, Tu K. Identifying cases of congestive heart failure from administrative data: a validation study using primary care patient records. Chronic diseases and injuries in Canada 2013; 33: 160-6. | **Database**<br>Ontario Congestive Heart Failure Database (CHF)<br><br>(March 31, 2019) |

| Chronic Condition | Case Definition/Reference | Database (latest data available) / *Notes |
|---|---|---|
| Diabetes | **Case Definition**<br>≥1 hospitalization (from DAD) or ≥2 physician billings with a diabetes fee code within a two-year period or ≥1 diabetes medication prescription<br><br><u>OHIP</u><br>OHIP physician diagnostic code: 250<br>OHIP physician service codes: Q040, K029, K030, K045, K046<br><br><u>DAD</u><br>ICD-9 diagnostic code: 250<br>ICD-10 diagnostic codes: E10, E11, E13, E14<br><br>**Reference**<br>Hux JE, Ivis F, Flintoft V, Bica A. Diabetes in Ontario Determination of prevalence and incidence using a validated administrative data algorithm. Diabetes care 2002; 25: 512-6. | **Database**<br>Ontario Diabetes Database (ODD)<br><br>(March 31, 2019) |
| Hypertension | **Case Definition**<br>≥1 hospitalization (from DAD) or ≥2 physician billings (from OHIP) with a hypertension fee code within a two-year period<br><br><u>OHIP</u>:<br>OHIP physician diagnostic codes: 401, 402, 403, 404, or 405<br><br><u>DAD, SDS</u>:<br>ICD-9 diagnostic codes: 401, 402, 403, 404, 405<br>ICD-10 diagnostic codes: I10, I11, I12, I13, I15<br><br>**Reference**<br>Tu K, Campbell NR, Chen Z-L, Cauch-Dudek KJ, McAlister FA. Accuracy of administrative databases in identifying patients with hypertension. Open Medicine 2007; 1: 18-26. | **Database**<br>Ontario Hypertension Database (HYPER)<br><br>(March 31, 2019)<br><br>*The HYPER database is limited to adults age 20 and older. |

Abbreviations: OHIP, Ontario Health Insurance Plan; DAD, Discharge Abstract Database; NACRS, National Ambulatory Care Reporting System Database; SDS, Same Day Surgery Database; OMHRS, Ontario Mental Health Reporting System,

Note: These chronic condition databases were created at ICES using validated case-finding algorithms to identify individuals with specific conditions in the health administrative databases.

### Table A.2. Definitions of Census-Based Neighbourhood Characteristics: constructs, statistical units, and operational definitions, using the 2016 (Ontario-specific) Canadian census data

| Construct | Statistical Unit | Operational Definition (Unit) | Census Variables used in the Ontario Census Area Profile (dataset) |
|---|---|---|---|
| **(A) Occupation Type** | | | |
| Percentage of people in managerial / professional Jobs | Persons | Percentage of total labour force population, age 15 and older, in executive, managerial, or professional occupations, based on the National Occupational Classification 2016 | F61 / F60 (Labour) |
| Percentage of people in sales / service Jobs | Persons | Percentage of total labour force population, age 15 and older, in sales and service occupations, based on the National Occupational Classification 2016 | F67 / F60 (Labour) |
| **(B) Transportation** | | | |
| Percentage of households driving | Private Household | Percentage of households driving a car, truck, or van as the main mode of commuting for the employed labour force, age 15 and older | F197 / F196 (Labour) |
| **(C) Housing** | | | |
| Average number of household occupants | Private Household | Average number of persons per private household | F18 (Family) |
| **(D) Income** | | | |
| Median after-tax income in 2015 | Private Household | Median after-tax income in 2015 among recipients for the population, aged 15 and older | F5 (Income) |

Note: Private household excludes people living outside of Canada and people living in collective dwellings (e.g., hospitals, nursing homes, penitentiaries, student residences). When the statistical unit is persons, it includes people in the total population.

The neighbourhood was defined according to forward-sortation areas (FSA). In Canada, the FSA represents a geographic region that starts with the same three letter postal code. In 2016, the province of Ontario, Canada, had 523 FSAs represented in the 2016 Census.
